# How do socioeconomic trajectories experienced during early adulthood contribute to the development of cardiometabolic health in young adults?

**DOI:** 10.1101/2024.09.02.24312850

**Authors:** Eleanor Winpenny, Jan Stochl, Alun Hughes, Kate Tilling, Laura D Howe

## Abstract

**Introduction:** Socioeconomic position has been strongly associated with cardiovascular health. However, little is known about the short-term health impacts of socioeconomic exposures during early adulthood. In this study we describe distinct socioeconomic trajectories of early adulthood (age 16-24y), and assess associations of these trajectories with measures of cardiometabolic health at age 24y.

**Methods:** Participants of the Avon Longitudinal Study of Parents and Children (ALSPAC), with data across age 16y to 24y (2007-2017) were included (n=7,568). Longitudinal latent class analysis identified socioeconomic trajectories, based on education and employment data across ages 16–24y. Cardiometabolic outcomes at age 24y comprised anthropometric, vascular, metabolic and cardiovascular structure and function measures. We modelled differences in cardiometabolic outcomes at age 24y across the socioeconomic trajectory classes, adjusting for childhood socioeconomic position, adolescent health behaviours and adolescent health.

**Results:** Four early adulthood socioeconomic trajectories were identified: (1) Higher Education (41% of the population), (2) Extended Education (9%), (3) Part-Time Employment (21%), and (4) Early Employment (29%). Associations between socioeconomic trajectory and cardiometabolic outcomes differed by sex. Among males, the Higher Education and Extended Education classes showed a healthier cardiometabolic profile, and the Part-time Employment class the least healthy. Among females there was less clear distinction between the classes, and the pattern across different outcomes was not consistent.

**Conclusion:** The newly identified ‘Part-time Employment’ class showed the least healthy cardiometabolic profile, and further research should focus on this group to understand the exposures contributing to poor cardiometabolic health in this sector of the population.

## Introduction

A large number of studies have highlighted differences in cardiovascular disease (CVD) according to socioeconomic position (SEP),[1–3] however we still do not have a detailed understanding of what is driving these inequalities or how best to address them. Longitudinal cohort studies have suggested that the impacts of socioeconomic factors on cardiovascular outcomes may accumulate across the life course, incorporating both childhood and adulthood SEP [4–7].

Early adulthood (age 16-24) is an important time of social and economic transition [8], when time spent in education, different types of employment, or not in education, employment or training (NEET) will contribute to the development of individual’s adult SEP [9]. Over recent decades, the school-to-work transition has become more protracted and complex, with greater diversity of transition pathways [10]. In particular, a greater need for higher education has led to many young people simultaneously participating in both education and employment [11]. However, much of the analysis of the role of early adulthood SEP in health inequalities remains focused on years of education or completion of a higher degree, not taking into account the diverse socioeconomic trajectories found in contemporary cohorts [8].

Early adulthood is also an important time for development of cardiovascular health; this is the age range when overweight and obesity increases the fastest in contemporary cohorts [12] and a period of rapid increase in prevalence of atherosclerotic lesions [13]. Previous research has suggested that early adulthood may be an important period for the development of socioeconomic inequalities in later cardiovascular health. Research using data from the 1970 British Cohort Study showed associations between different trajectories of SEP across early adulthood with a range of cardiometabolic risk factors in mid-adulthood (age 46), independent of childhood SEP, and not mediated by later adulthood SEP [14]. There is evidence that transitions between education and employment in early adulthood contribute to changes in behavioural cardiovascular risk factors over this period, for example diet [15], physical activity [16], smoking and alcohol intake [17], which may provide mechanisms contributing to the development of cardiovascular inequalities across early adulthood.

However, it is currently unknown whether cardiometabolic inequalities related to young adults’ own socioeconomic position can already be identified in young adulthood. A wide range of anthropometric and blood-based biomarkers, associated with cardiovascular outcomes later in life, have previously shown associations with parental SEP [18]. However, cardiovascular risk factors develop at different rates by sex, with young women showing better lipid profiles than men [19,20], and lower blood pressure [21]. More recently, structural measures of the heart and vasculature, which show independent associations with cardiovascular outcomes [22], have become available in cohorts. Carotid intima-media thickness (c-IMT) is a measure of the extent of atherosclerosis development in the carotid artery, pulse-wave velocity is a measure of arterial stiffness, while left ventricular mass index and relative wall thickness represent measures of the structure of the heart. c-IMT and PWV have previously shown associations with parental SEP in adolescence [23–25], however it is not known how quickly these risk factors may change in response to early adulthood exposures.

In this study we aimed to examine the contribution of trajectories of SEP in early adulthood to a wide range of measures of cardiometabolic health in early adulthood, addressing three questions:

**Q1: What are the socioeconomic trajectories followed by different classes of the population across early adulthood (age 16-24y)?**

**Q2: What are the demographic characteristics of the population making up each socioeconomic trajectory classes?**

**Q3: How is socioeconomic trajectory class during early adulthood associated with cardiovascular health in males and females at age 24, and are these associations independent of childhood SEP prior to age 16?**

## Methods

### Survey design and participants

The Avon Longitudinal Study of Parents and Children (ALSPAC) is a large, prospective cohort study based in the South West of England, aimed at investigating the environmental and genetic influences on the health, behaviour and development of those born in the early 1990s. All pregnancies in the former Avon Health Authority with an expected delivery date between 1 April 1991 and 31 December 1992 were eligible for the study; 14,541 pregnancies were initially enrolled, resulting in a cohort of 14,062 live births and 13,988 children who were alive at 1 year of age [26,27]. The social and demographic characteristics of the women enrolled were similar to those found in the 1991 National Census survey of Great Britain, although mothers were more likely than those across Great Britain to be White, to live in owner-occupied accommodation and to have a car available to the household [28].

Study data were collected and managed using REDCap electronic data capture tools hosted at the University of Bristol [29]. REDCap (Research Electronic Data Capture) is a secure, web-based software platform designed to support data capture for research studies.

Ethical approval for the study was obtained from the ALSPAC Law and Ethics Committee and the Local Research Ethics Committees, and conformed to the Declaration of Helsinki. Consent for biological samples has been collected in accordance with the Human Tissue Act (2004). Informed consent for the use of data collected via questionnaires and clinics was obtained from participants following the recommendations of the ALSPAC Ethics and Law Committee at the time. If the child was younger than age 16 years, informed written consent was obtained from the parent/guardian alongside assent from the child. When participants were age 16 years or older, they provided their own informed written consent.

### Exposure: early adulthood socioeconomic position

Education and employment data were self-reported at ages 16, 18, 20, 21, 22, 23, 24y, from 2007 to 2017. Education data included whether the participant was (1) in education full time, (2) in education part time, or (3) not in education/training, at all time points except age 16, when only full time education or not in education/training were recorded. Employment data included data on whether participants were (1) in full-time employment, (2) in part-time employment, (3) were unemployed and looking for work, or (4) were not working due to education participation, looking after home/family or another reason. At ages 20, 21, 22 and 23y income data was also available, which was collapsed into 3 categories, (low income) those earning less than £1000/month, (mid-income) between £1000-1500/month and (high income) greater than £1500 per month. At ages 20y and 21y, a low income cut-off of £900/month was used instead of £1000/month, as the categories provided in the questionnaire had slightly different boundaries. Employment and income data were combined, such that those in full-time employment could be categorised by level of income. This resulted in 7 employment categories: Full time employment (high income), full-time employment (mid-income), full-time employment (low-income), full-time employment (income unknown) part-time employment, unemployed, not working.

Occupational social class was measured for those working at age 23, and classified according to the NS-SEC categories. Educational level attained by age 24y was derived from data collected at age 26y on qualifications completed and year in which they were obtained. The highest level of qualification obtained by age 24 was categorised into the following categories: Level 1/2 vocational, GCSE, Level 3 vocational, A-level, Level 4/5 vocational, Degree, Higher degree.

### Outcomes: cardiometabolic health

Data on cardiometabolic health at age 24y was collected at the F24 clinic session, which included anthropometric, blood pressure and heart rate measurements, DXA scans, blood assays, and among a subsample of participants, echocardiography, carotid intima-mediate thickness and pulse wave velocity measurement. Further details on all measurements can be found in the ALSPAC data dictionary, available on the ALSPAC website (http://www.bristol.ac.uk/alspac/).

From these measurements, body fat percentage was calculated as total body fat mass as a percentage of total body mass. Glucose and insulin measures were used to calculate HOMA-IR, using the HOMA2 Calculator software provided by the Diabetes Trials Unit, University of Oxford [30]. Measures of triglycerides, HOMA-IR and CRP were log-transformed, as these variables were positively skewed. We excluded those participants with values of CRP above 10mg/L from CRP analyses, as this is likely to reflect recent infection rather than chronic inflammation. Relative wall thickness (RWT), defined as 2 times the posterior wall thickness divided by the left ventricular (LV) diastolic diameter and left ventricular mass indexed to height^1.7 (LVMI) were computed as indicators of cardiac structure [31].

### Descriptive variables and covariates

Childhood sociodemographic variables were collected by parental questionnaire. We included childhood sociodemographic variables collected at age 4, age 8, and age 16 as descriptive variables with choice of age based on level of missing data, and quality of the data collected (ie interpretability of categories used). Individual data on partnership and parenthood were self-reported at age 24y

In addition to sex and ethnicity, three groups of potential confounders were included in the analysis: (1) childhood SEP (mother’s age during pregnancy, parity, maternal education at age 16, neighbourhood deprivation (Index of Multiple Deprivation) at age 16, family structure at age 8), (2) adolescent health behaviours at age 16 (alcohol intake, smoking, cannabis use), (3) adolescent health measures (self-rated health 16y, depressive symptoms 16y, systolic blood pressure 15y, and waist circumference age 15y). We also tested inclusion of paternal occupational class and family income as childhood SEP covariates, but these variables were redundant in the model, making no difference to the results, so were removed.

### Statistical analysis

Latent class analysis was conducted using Mplus V8.7, to generate classes of individuals based on categorical data on education, employment and income (as described above) from age 16 to 24 years, their occupational social class at age 23y, and their educational attainment by age 24y. From here onwards we refer to these classes as ‘socioeconomic trajectory classes’. All individuals who had any data on these variables from age 16 to 24 years were included (n=7,568). The latent class analysis was adjusted for age, to take account of the variation in age of participants within each wave of data. Classes were added to the model sequentially, from one to six classes, beyond which the model failed to converge on a stable solution. The final number of latent classes used for the analysis was selected based on fit statistics (Akaike information criterion, Bayesian information criterion, Vuong-Lo-Mendell-Rubin likelihood ratio test, bootstrapped parametric likelihood ratio test) together with the interpretability of the classes generated [32]. Entropy was reported, but not used in model selection. Missing data on latent class indicators were addressed using full information maximum likelihood estimation.

Descriptive analyses and multiple imputation of covariates were performed using Stata, V.16. Descriptive analyses of class membership compared descriptive sociodemographic variables and cardiometabolic outcome variables across classes. We used chi-squared tests, ANOVA or Kruskal-Wallis tests (for non-normal outcomes) to test for equal distribution of sociodemographic variables across the four classes.

Missing covariate data were imputed by chained equations (under the missing at random assumption) using the Stata ‘mi impute chained’ command. To impute covariates, a single imputation model was used for each sex, which included the exposure, all outcomes, covariates and auxiliary variables. We included additional variables on parental SEP from various childhood waves (education level, socioeconomic class, income and area level deprivation), as auxiliary variables, creating 100 imputed datasets (see online supplementary methods and supplementary table S1 for details). A single imputation model by sex was considered appropriate across all outcomes, as the additional outcomes could also be considered to be auxiliary variables. A comparison of those participating in the cohort at age 16 with those originally enrolled in ALSPAC and national data has been reported elsewhere [26].

Analysis of associations between SEP class and each cardiometabolic outcome were conducted in Mplus, using the manual BCH method to estimate a distal outcome model, as described in Asparouhov and Muthen (2021). This model uses weights to reflect the measurement error of the latent class variable, and allows estimation of the intercept of the outcome variable in each SEP class, adjusted for covariates. The Mplus code for association of SEP class and outcomes can be found in the Supplementary Methods. Based on previous evidence for sex differences in the relationship between SEP and cardiovascular disease, all analyses were performed stratified by sex. Participants were excluded from the outcome analysis if they were missing data on sex (n=11), or reported that they were pregnant (n=19) or might be pregnant (n=12) at age 24y, leaving up to n=7526 participants for the outcome analysis. Coded data was not available on medications related to the outcomes at age 24, but based on data from age 17, these numbers are likely to be very low and unlikely to influence the results. We also did not have data in our dataset on pre-existing cardiometabolic disease, but again numbers are low (<50) and unlikely to influence findings. Associations between SEP class and each cardiometabolic outcome was analysed separately, and only participants with data on this outcome were included in each analysis. P-values were not adjusted for multiple testing, based on recommendations [34], since our outcomes are not independent, and to allow interpretation of variance parameters by the reader, avoiding interpretation based on statistical significance.

## Results

### Early adulthood socioeconomic trajectories

Sequential addition of classes to the latent class analysis found that while some fit statistics (AIC, BIC) continued to improve with up to six classes, this was at a decreasing rate (see online supplemental table S2). The Vuong-Lo-Mendell-Rubin Likelihood Ratio Test suggested a 4-class model fitted best, and examination of the response probabilities for the indicator variables suggested that this model presented a solution that was interpretable as distinct population groups (Figure 1). Addition of a 5^th^ class appeared to further split the Higher Education class into 2 fairly similar classes, and we therefore returned to the 4-class solution. As shown in Figure 1, this included (1) a Higher Education class (41%), who largely remained in education to around age 21y, many obtaining degree-level or above, and then entered full-time employment, (2) an Extended Education class, the majority of whom remained in full-time education up to age 24y, some attaining higher degrees but including 40% attaining only A-level qualifications or below by age 24y, (3) a Part-time Employment class who had mostly finished education after age 21, but were more likely to remain in part-time employment and (4) an Early Employment class most of whom finished full-time education around age 18y and entered full-time employment. Figure 1 shows each class represented in a column, with graphs representing the response probabilities for each of the education and employment variables seen in that class. Employment and education participation categories are shown across each year of age where data was available, from age 16 to 24 years, while occupational socio-economic classification and education level attained are presented at age 23 and 24 years respectively.

**Fig. 1.**
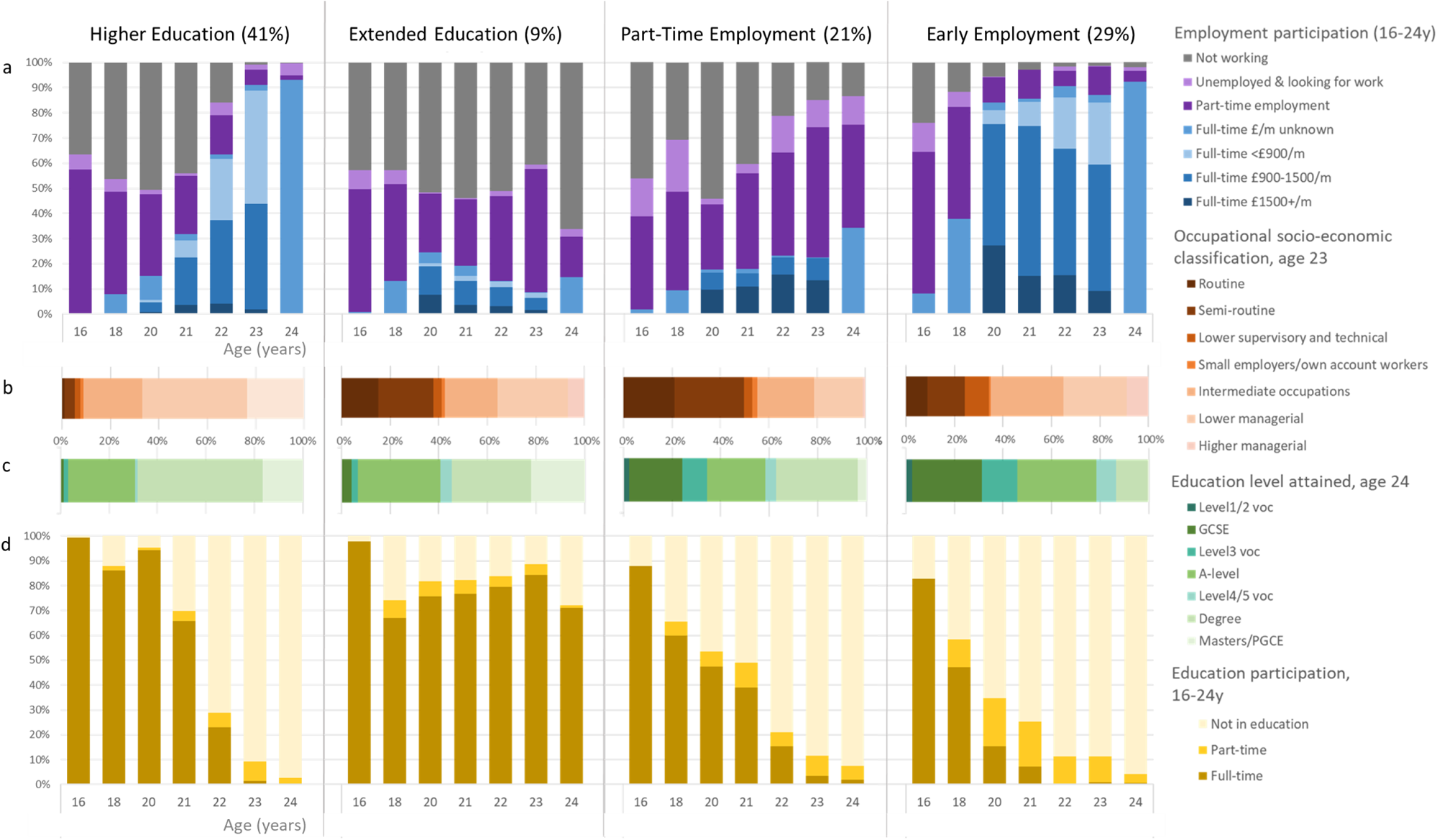
Education and employment participation and level, by socioeconomic trajectory class. Each column depicts data for a single socioeconomic trajectory class. Row (a) shows employment participation across ages 16-24 years, row (b) shows occupational socio-economic classification at age 23, row (c) shows education level attained by age 24y, row (d) shows education participation across ages 16-24 years.

Table 1 presents differences in sociodemographic data across the different SEP classes by sex, based on assigning the most likely class for each individual. There were differences in childhood SEP measures between the latent classes, with the Higher Education and Extended Education classes having indicators of higher childhood SEP than the Part-time Employment and Early Employment classes. The Extended Education and Part-time Employment classes also had a higher percentage of members who have a partner. Most of the patterns in sociodemographic variables observed across the different classes were fairly consistent between the sexes. Notably, there were a higher proportion of females in the Part-time Employment class, and women in this class were much more likely to have children by age 24y.

**Table 1:** Descriptive sociodemographic data by early adulthood SEP class.

|  | Males (n=3164) |  |  |  |  | Females (n=4393) |  |  |  |  |
| --- | --- | --- | --- | --- | --- | --- | --- | --- | --- | --- |
|  | Higher Education | Extended Education | Part-time Employment | Early Employment | p-value | Higher Education | Extended Education | Part-time Employment | Early Employment | p-value |
|  | 43% | 9% | 18% | 30% |  | 40% | 10% | 23% | 28% |  |
| Ethnicity, % white (n=6678) | 95.8 | 93.9 | 95.6 | 96.5 | 0.03 | 95.7 | 94.6 | 95 | 97 | 0.09 |
| Childhood covariates |  |  |  |  |  |  |  |  |  |  |
| Maternal age at gestation, years mean (SD) (n=6841) | 29.4 (4.5) | 29.9 (4.2) | 29.2 (4.9) | 28.1 (4.6) | 0.02 | 29.0 (4.4) | 29.8 (4.4) | 27.9 (4.8) | 27.5 (4.6) | 0.01 |
| Maternal education at 16y, % NVQ level 4 or higher (n=4392) | 48.7 | 58.3 | 44.7 | 27.5 | <0.001 | 48.4 | 56.5 | 38.3 | 23.3 | <0.001 |
| Father NS-SEC (5 class) at age 4y, % Higher managerial, administrative and professional occupations (n=4106) | 47.0 | 58.9 | 37.4 | 24.3 | <0.001 | 43.2 | 54.8 | 32.8 | 21.7 | <0.001 |
| IMD quintiles at age 16y, % in least deprived quintile (n=4927) | 43.8 | 42.9 | 34.4 | 33.0 | <0.001 | 40.9 | 36.9 | 29.4 | 32.1 | <0.001 |
| Family structure at age 8y, % single parent families (n=5669) | 8.1 | 7.2 | 9.2 | 7.2 | 0.63 | 7.3 | 9.8 | 13.1 | 10.7 | <0.001 |
| Adulthood covariates |  |  |  |  |  |  |  |  |  |  |
| Hours worked per week among those working at 23y, mean (SD) (n=3336) | 39.9 (12.0) | 23.3 (17.5) | 30.0 (15.0) | 38.1 (14.5) | <0.001 | 40.5 (13.1) | 22.2 (16.9) | 28.1 (15.0) | 36.7 (12.3) | <0.001 |
| Take-home pay among those working, age 23y, % less than £1000/month (n=3465) | 4.5 | 74.7 | 71.6 | 10.3 | <0.001 | 3.9 | 78.1 | 83.2 | 16.3 | <0.001 |
| Partnership status, % with partner at age 24y (n=5094) | 38.3 | 45.9 | 40.1 | 33 | <0.001 | 30.5 | 35.3 | 40.1 | 31.2 | <0.001 |
| Family status, % a parent by age 24y (n=6309) | 3.1 | 1.5 | 6.5 | 10.1 | <0.001 | 3.22 | 6.44 | 32.9 | 13.3 | <0.001 |

### Associations between socioeconomic trajectory class and cardiometabolic risk factors

Adjusted analysis of the association of early adulthood socioeconomic trajectory class with cardiometabolic outcomes are presented in Table 2, and standardised results depicted in Figure 2 for comparison across different outcomes. Unadjusted results are shown in Supplementary Table S3. Across many outcomes, the pattern of associations was very similar between adjusted and unadjusted outcomes, although in many cases the difference between classes was slightly attenuated after adjustment. In a small number of cases the differences between classes were greater after adjustment for confounders.

**Fig. 2.**
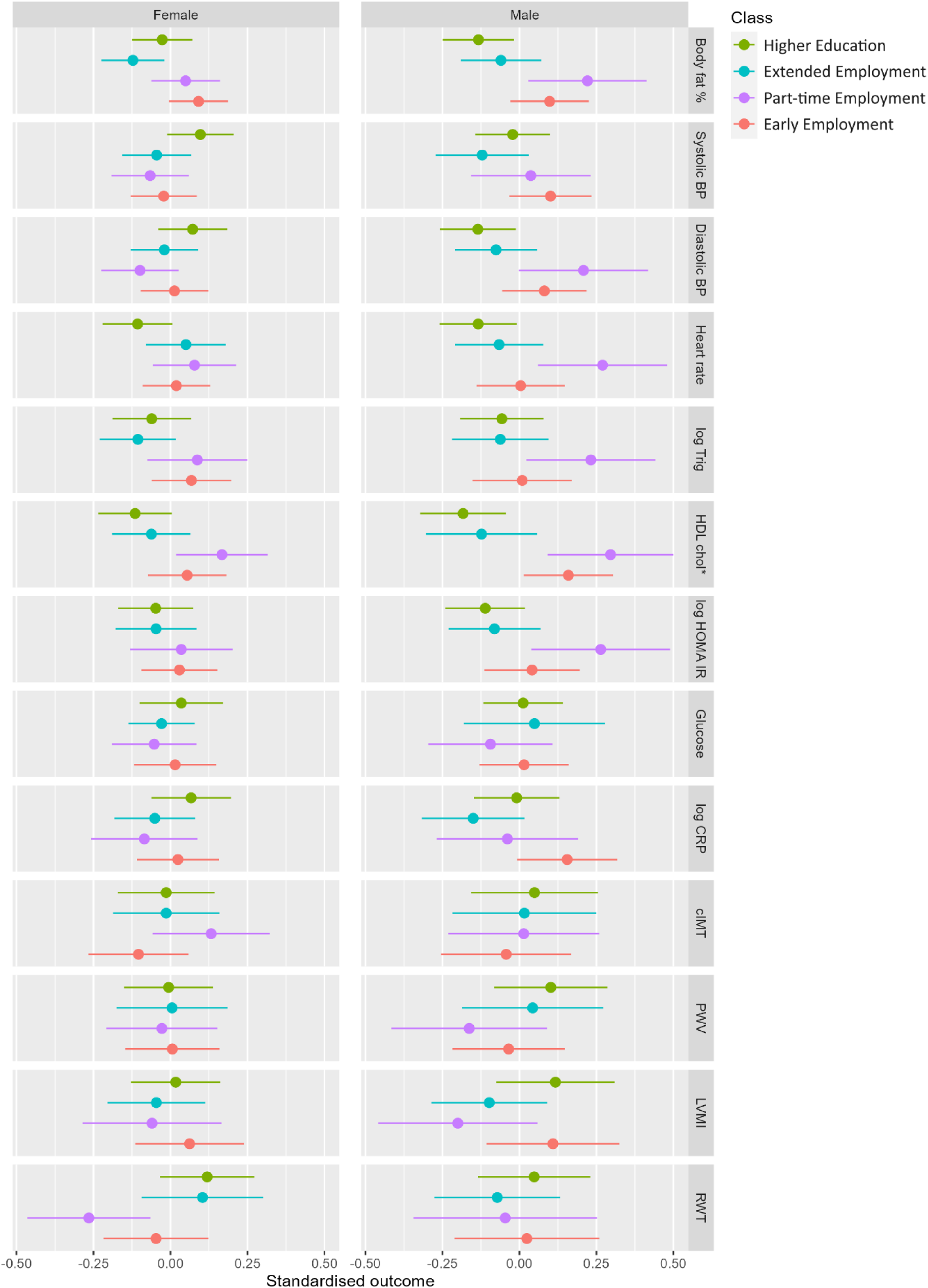
Standardised estimated means and 95% confidence intervals of each outcome for each socioeconomic trajectory class, adjusted for covariates. Legend: Green=Higher Education, Cyan = Extended Education, Purple=Part-time Employment, Red=Early Employment. *HDL cholesterol results multiplied by −1, in order that across all outcomes positive values are detrimental for health

**Table 2:**
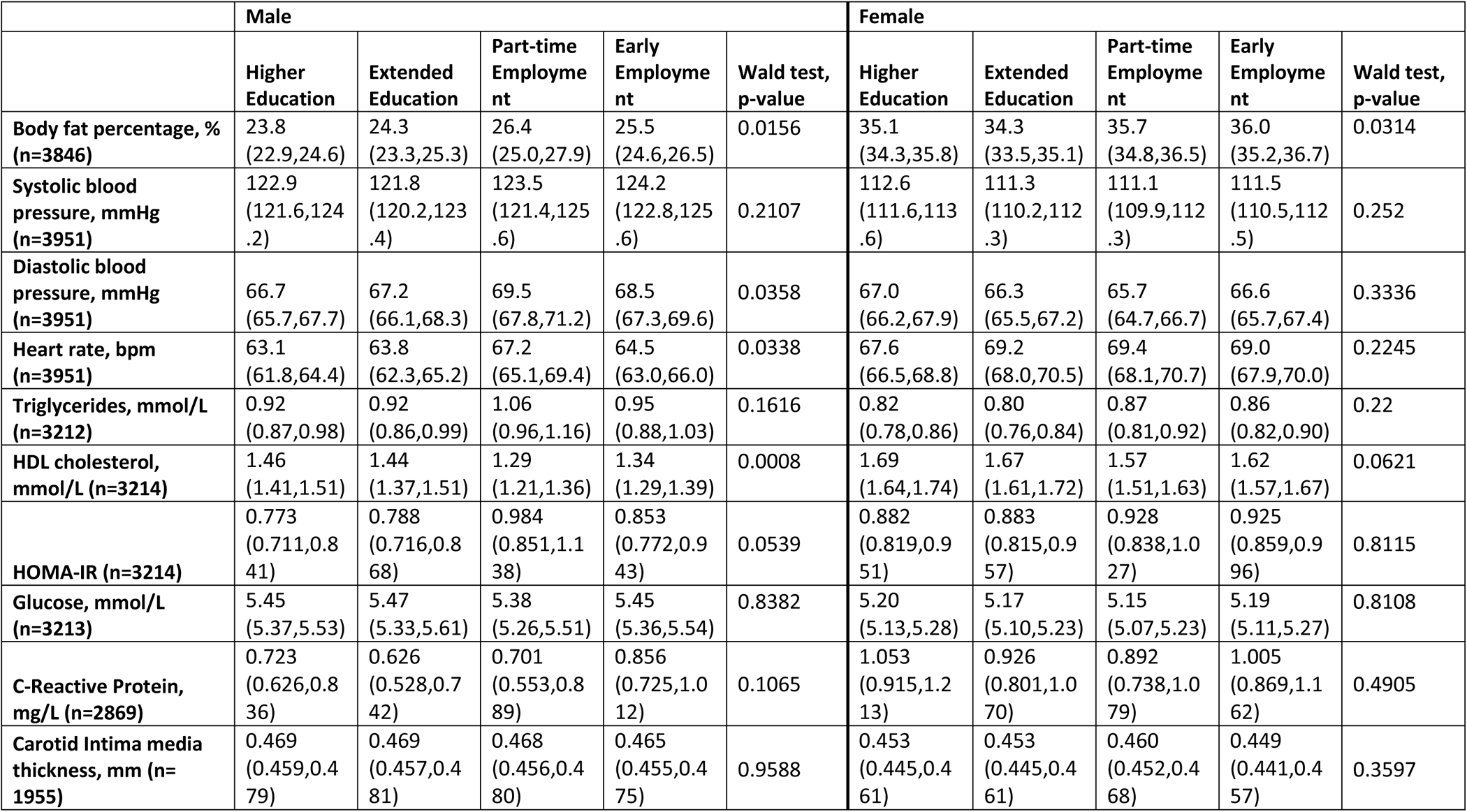
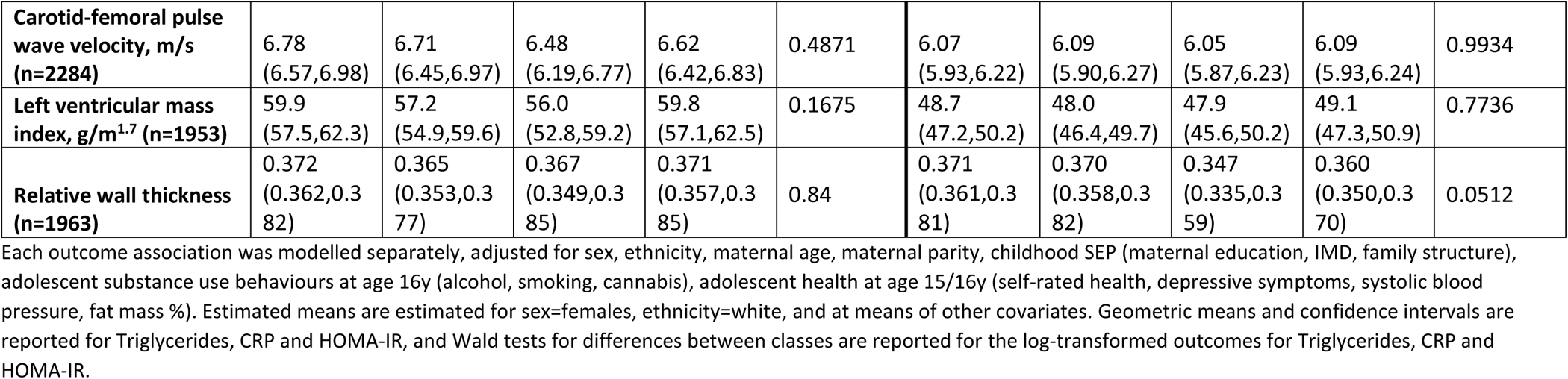
Estimated means (with 95% confidence intervals) of each outcome for each socioeconomic trajectory class, adjusted for covariates.

|  | Male |  |  |  |  | Female |  |  |  |  |
| --- | --- | --- | --- | --- | --- | --- | --- | --- | --- | --- |
|  | Higher Education | Extended Education | Part-time Employment | Early Employment | Wald test, p-value | Higher Education | Extended Education | Part-time Employment | Early Employment | Wald test, p-value |
| **Body fat percentage, % (n=3846)** | 23.8<br>(22.9,24.6) | 24.3<br>(23.3,25.3) | 26.4<br>(25.0,27.9) | 25.5<br>(24.6,26.5) | 0.0156 | 35.1<br>(34.3,35.8) | 34.3<br>(33.5,35.1) | 35.7<br>(34.8,36.5) | 36.0<br>(35.2,36.7) | 0.0314 |
| **Systolic blood pressure, mmHg (n=3951)** | 122.9<br>(121.6,124.2) | 121.8<br>(120.2,123.4) | 123.5<br>(121.4,125.6) | 124.2<br>(122.8,125.6) | 0.2107 | 112.6<br>(111.6,113.6) | 111.3<br>(110.2,112.3) | 111.1<br>(109.9,112.3) | 111.5<br>(110.5,112.5) | 0.252 |
| **Diastolic blood pressure, mmHg (n=3951)** | 66.7<br>(65.7,67.7) | 67.2<br>(66.1,68.3) | 69.5<br>(67.8,71.2) | 68.5<br>(67.3,69.6) | 0.0358 | 67.0<br>(66.2,67.9) | 66.3<br>(65.5,67.2) | 65.7<br>(64.7,66.7) | 66.6<br>(65.7,67.4) | 0.3336 |
| **Heart rate, bpm (n=3951)** | 63.1<br>(61.8,64.4) | 63.8<br>(62.3,65.2) | 67.2<br>(65.1,69.4) | 64.5<br>(63.0,66.0) | 0.0338 | 67.6<br>(66.5,68.8) | 69.2<br>(68.0,70.5) | 69.4<br>(68.1,70.7) | 69.0<br>(67.9,70.0) | 0.2245 |
| **Triglycerides, mmol/L (n=3212)** | 0.92<br>(0.87,0.98) | 0.92<br>(0.86,0.99) | 1.06<br>(0.96,1.16) | 0.95<br>(0.88,1.03) | 0.1616 | 0.82<br>(0.78,0.86) | 0.80<br>(0.76,0.84) | 0.87<br>(0.81,0.92) | 0.86<br>(0.82,0.90) | 0.22 |
| **HDL cholesterol, mmol/L (n=3214)** | 1.46<br>(1.41,1.51) | 1.44<br>(1.37,1.51) | 1.29<br>(1.21,1.36) | 1.34<br>(1.29,1.39) | 0.0008 | 1.69<br>(1.64,1.74) | 1.67<br>(1.61,1.72) | 1.57<br>(1.51,1.63) | 1.62<br>(1.57,1.67) | 0.0621 |
| **HOMA-IR (n=3214)** | 0.773<br>(0.711,0.841) | 0.788<br>(0.716,0.868) | 0.984<br>(0.851,1.138) | 0.853<br>(0.772,0.943) | 0.0539 | 0.882<br>(0.819,0.951) | 0.883<br>(0.815,0.957) | 0.928<br>(0.838,1.027) | 0.925<br>(0.859,0.996) | 0.8115 |
| **Glucose, mmol/L (n=3213)** | 5.45<br>(5.37,5.53) | 5.47<br>(5.33,5.61) | 5.38<br>(5.26,5.51) | 5.45<br>(5.36,5.54) | 0.8382 | 5.20<br>(5.13,5.28) | 5.17<br>(5.10,5.23) | 5.15<br>(5.07,5.23) | 5.19<br>(5.11,5.27) | 0.8108 |
| **C-Reactive Protein, mg/L (n=2869)** | 0.723<br>(0.626,0.836) | 0.626<br>(0.528,0.742) | 0.701<br>(0.553,0.889) | 0.856<br>(0.725,1.012) | 0.1065 | 1.053<br>(0.915,1.213) | 0.926<br>(0.801,1.070) | 0.892<br>(0.738,1.079) | 1.005<br>(0.869,1.162) | 0.4905 |
| **Carotid Intima media thickness, mm (n=1955)** | 0.469<br>(0.459,0.479) | 0.469<br>(0.457,0.481) | 0.468<br>(0.456,0.480) | 0.465<br>(0.455,0.475) | 0.9588 | 0.453<br>(0.445,0.461) | 0.453<br>(0.445,0.461) | 0.460<br>(0.452,0.468) | 0.449<br>(0.441,0.457) | 0.3597 |
| **Carotid-femoral pulse wave velocity, m/s (n=2284)** | 6.78<br>(6.57,6.98) | 6.71<br>(6.45,6.97) | 6.48<br>(6.19,6.77) | 6.62<br>(6.42,6.83) | 0.4871 | 6.07<br>(5.93,6.22) | 6.09<br>(5.90,6.27) | 6.05<br>(5.87,6.23) | 6.09<br>(5.93,6.24) | 0.9934 |
| **Left ventricular mass index, g/m<sup>1.7</sup> (n=1953)** | 59.9<br>(57.5,62.3) | 57.2<br>(54.9,59.6) | 56.0<br>(52.8,59.2) | 59.8<br>(57.1,62.5) | 0.1675 | 48.7<br>(47.2,50.2) | 48.0<br>(46.4,49.7) | 47.9<br>(45.6,50.2) | 49.1<br>(47.3,50.9) | 0.7736 |
| **Relative wall thickness (n=1963)** | 0.372<br>(0.362,0.382) | 0.365<br>(0.353,0.377) | 0.367<br>(0.349,0.385) | 0.371<br>(0.357,0.385) | 0.84 | 0.371<br>(0.361,0.381) | 0.370<br>(0.358,0.382) | 0.347<br>(0.335,0.359) | 0.360<br>(0.350,0.370) | 0.0512 |

Each outcome association was modelled separately, adjusted for sex, ethnicity, maternal age, maternal parity, childhood SEP (maternal education, IMD, family structure), adolescent substance use behaviours at age 16y (alcohol, smoking, cannabis), adolescent health at age 15/16y (self-rated health, depressive symptoms, systolic blood pressure, fat mass %). Estimated means are estimated for sex=females, ethnicity=white, and at means of other covariates. Geometric means and confidence intervals are reported for Triglycerides, CRP and HOMA-IR, and Wald tests for differences between classes are reported for the log-transformed outcomes for Triglycerides, CRP and HOMA-IR.

In general, stronger associations between socioeconomic trajectory and cardiometabolic outcomes were seen for males compared to females. Among males, differences between the classes could be observed for body fat percentage, diastolic blood pressure, heart rate, HDL cholesterol, and insulin resistance, although the effect sizes were fairly small at age 24y. For example, estimated mean body fat percentage was 23.8% (95%CI 22.9,24.6) in the Higher Education class, but 25.5% (95%CI 24.6,26.5) in the Early employment class, and 26.4% (95% CI 25.0,27.9) in the Part-time employment class. Similarly diastolic blood pressure was lowest at 66.7mmHg (95%CI 65.7,67.7) in the Higher Education class, and highest at 69.5mmHg (95%CI 67.8,71.2) in the Part-time Employment class. Across these outcomes, a similar pattern emerged with the ‘Part-time Employment’ class consistently showing features indicative of higher cardiometabolic risk, followed by the Early Employment class. The ‘Higher education’ and ‘Extended Education’ classes had indicators of better cardiovascular health across these outcomes. Differences in cardiometabolic outcomes between socioeconomic trajectory classes in females were much less consistent, and were imprecisely estimated. Among women, relative wall thickness, a measure of left ventricular remodelling, was lower in Part-Time Employment class, compared to other classes. One suggestion was that this may be due to the effect of previous pregnancy, however a sensitivity analysis restricted to females without children found almost identical results to the main analysis (Supplementary Table S4).

## Discussion

This study explored the socioeconomic trajectories of young adults aged 16-24, over the years 2007-2017. Our analysis revealed 4 distinct socioeconomic trajectories of early adulthood: a large Higher Education trajectory (41%) who mostly attained university-level qualifications and entered managerial employment, an Early Employment trajectory of those who left education and entered employment around age 18 (29%), a Part-time Employment trajectory of those who continued in part-time employment across early adulthood (21%), and a small Extended Education trajectory who remained in education across early adulthood up to age 24y (9%).

Associations between socioeconomic trajectories and cardiometabolic health varied by sex. Both males and females showed associations between socioeconomic trajectory and body fat percentage, after adjustment for parental SEP and other covariates. However, clear differences in diastolic blood pressure, heart rate, HDL cholesterol and insulin resistance across socioeconomic trajectory classes were seen only among males. In general in males the Higher Education and Extended Education classes showed a healthier profile, with the Early Employment and Part-time Employment classes less healthy. Differences between the remaining outcomes were imprecisely estimated, although consistent patterns could be seen across a number of outcomes, which may develop further with age.

### Comparison of identified socioeconomic trajectories with findings from other studies

The majority of previous studies exploring socioeconomic trajectories, or work-family trajectories, are based on cohort studies where individuals were born prior to 1980, resulting in early adulthood trajectories which do not reflect contemporary lives [35]. For example, our own analysis of socioeconomic trajectories from the 1970 British Cohort Study, using a similar methodology, identified six fairly simple trajectories, including a Continued Education trajectory comprising the 20% of the population who continued in education beyond age 18, five trajectories reflecting transitions into employment of different occupational social class, and one trajectory (8%) who became economically inactive [14]. Others have described a declining occurrence of a direct school-to-work transition from the 70s to the late 80s,[36] and the changing context in recent decades has resulted in a very different experience of becoming an adult [8], and it is no surprise that the trajectories observed in this study are very different to those observed previously.

We identified two studies of more recent UK data, one making use of a subset of the Understanding Society dataset including those born from 1985-89, and another of the Longitudinal Study of Young People in England. These used sequence analysis to identify school-work trajectories, reporting on five and six trajectory clusters respectively [8,37]. Several of these clusters overlapped with the classes identified in this study, with a similar size Higher Education and Early Employment group identified in the LSYPE analysis, a cohort which is contemporary with the ALSPAC cohort [37]. However in both cases classes defined by unemployment or NEET were observed, which we did not see in our study.

Instead, since we were able to differentiate between full-time and part-time employment, we identified a ‘Part-time employment’ class, representing 21% of the study population. The majority of this group were unemployed or in part-time employment across ages 16-24y, with 60% attaining lower than degree-level education by age 24y. The existence of this group may reflect recent changes in the employment landscape. Others have documented a sharp increase in underemployment, particularly among young workers, following the 2008 UK recession, which persisted for much of the next decade. This period aligns with the timing of early adulthood in the ALSPAC cohort [38,39]. Underemployment is defined as those who work part-time because there are insufficient full-time jobs available, and those who would prefer to work more hours [38]. Almost a quarter of our ‘Part-time employment’ class report children by age 24y, so for some it may be that part-time work is a choice rather than a necessity.

### Comparison of associations with cardiovascular and metabolic outcomes with previous findings

We found some evidence of greater inequality in cardiometabolic outcomes across the socioeconomic trajectory classes among men than women. Previous studies have reported on sex-specific differences in cardiovascular outcomes [40], however, few studies have focussed on cardiometabolic risk factors in young adults. Nevertheless, there is some literature to suggest that socioeconomic inequalities in cardiometabolic risk factors develop with age, such that greater inequalities are observed among young men, which subsequently decrease with age, while in women socioeconomic inequalities increase with age [41]. This would support our finding of greater differences among men at age 24 years, and we might expect to see inequalities in women developing later in life.

Our findings of lower body fat, and more favourable heart rate, diastolic blood pressure, HDL cholesterol and insulin resistance profiles among men in the Higher Education class are consistent with previous evidence of the relationships between SEP and cardiovascular health. Indeed, recent Mendelian randomisation studies have supported a causal link between years of education and cardiovascular disease [42,43], building on previous observational evidence. Our finding that these differences between the Higher Education group and Early Employment group can already be identified by age 24y (after adjusting for parental SEP) imply that cardiovascular inequalities related to individual education attainment are already beginning to accumulate during early adulthood, in response to environments experienced and behaviours sustained during this period. Although many of the outcomes showed similar patterns, some differences between classes were attenuated following adjustment, suggesting that both childhood and early adulthood SEP contribute to differences between classes in early adulthood. In a small number of cases (e.g. higher diastolic blood pressure and insulin resistance among males of the Part-time Employment class), differences between classes only emerged in the adjusted analysis, suggesting that childhood and early adulthood exposures may exert influences in different directions. More research is needed into the detail of differences in exposures over time in these population classes to understand the causes of the differential decline in cardiovascular health and the potential for intervention.

Our analyses found that the ‘Extended Education’ group also showed a favourable metabolic profile in men, with many cardiometabolic variables, e.g. body fat, blood pressure, HDL cholesterol and insulin resistance, being similar to the ‘Higher Education’ class. This group showed lower attainment of degree-level education by age 24y compared to the Higher Education class, suggesting delayed education rather than study towards higher degrees, for the most part. Nevertheless continued exposure to the educational (rather than workplace) environment appeared favourable for the outcomes studied. By contrast, the new ‘Part-time Employment’ class showed in general an unfavourable metabolic profile, with many metabolic outcomes e.g. body fat %, diastolic blood pressure, triglycerides, HOMA-IR appearing even less healthy than those of the Early Employment class. In some cases (e.g. diastolic blood pressure, HOMA-IR) the difference between this class and other classes increased after adjustment for covariates, suggesting that early adulthood exposures of the ‘Part-time Employment’ class operated in a different direction to childhood confounders. Wide confidence intervals suggest diversity of experiences in this class, with further research is needed to explore this socioeconomic trajectory class in more detail.

We included several measures of cardiovascular structure and function in our analyses. Although previous studies have identified socioeconomic differences in these measures in adolescence and early adulthood, in relation to childhood SEP [25,44], it is likely that the time frame for development of structural variation is longer than was possible within the current analysis of early adulthood exposures on outcomes at age 24y. It will be interesting to see how individual SEP inequalities in these structural measures develop as the ALSPAC cohort age.

### Strengths and Limitations

This study makes use of contemporary data on young adults from the ALSPAC cohort study, conducting a detailed classification of socioeconomic trajectories of early adulthood and relating these trajectories to a wide range of metabolic and cardiovascular outcomes. Classification of the population into a small number of socioeconomic trajectory classes allows us to explore population heterogeneity of socioeconomic trajectories and describe those commonly followed by the different sectors of the population [32]. It also allows a person-centred approach to analysis of associations with subsequent cardiometabolic health, taking into account the role of several aspects of SEP which change over time. Nevertheless, we acknowledge that the indicators we have included cannot capture all aspects of SEP, in particular indicators of wealth, or SEP at the level of the household rather than the individual [45]. In addition, the derived classes are not homogenous, such that there will always be some challenges in interpreting these classes, and differences in cardiometabolic outcomes between classes. For example, in this study the ‘Part-time Employment’ class included those working part-time for different reasons which might include family reasons, as well as unavailability of full-time jobs. Similarly those in ‘Extended Education’ include both those pursuing higher degrees and those who may have moved through education more slowly. This method of analysis therefore allows us to focus on the exposures experienced by each (e.g. degree of exposure to the educational or workplace environment) but cannot unpick the different reasons underlying such exposures.

The ALSPAC cohort is a large birth cohort, drawing on the population born in the Avon Health Authority in the South West of England. Although over 70% of the eligible pregnancies in the region were enrolled in the study, differential recruitment and attrition of this cohort compared to a national population sample (of England) means that by late adolescence the ALSPAC cohort sample included a higher proportion of female participants, showed higher white ethnicity, and higher household income compared to both the original sample population and a national population sample [26]. Our latent class method allowed us to include individuals who had reported on any of the education or employment measures included at any time-point during early adulthood (age 16-24y), making full use of all data available to derive socioeconomic trajectories. However, the early adulthood socioeconomic trajectories developed in our analysis are nevertheless based on this sample population, and may not be generalizable to a wider population sample. In analysis of the associations between socioeconomic trajectories and cardiometabolic health, we adjusted for a wide range of covariates, including sex, ethnicity and socioeconomic status, so that these observed characteristics are unlikely to confound our associations. However, unobserved characteristics related to participant attrition, such as unobserved differences in health status may have biased our reported associations. In this case, since we would expect those in worse cardiometabolic health, as well as those of lower SEP, to be more likely to drop out of the study [46] this may result in underestimation of relationships between social disadvantage and health, although based on previous studies we expect any bias introduced to be fairly limited [46,47].

Others have discussed that health selection may play an important role in accounting for socioeconomic inequalities in health [48]. In this analysis we were interested in the causal effect of socioeconomic trajectories on cardiometabolic health. We therefore tried as far as possible to adjust for potential confounding influences from adolescent behaviours and adolescent physical and mental health, as well as from childhood SEP, adjusting for a variety of parental SEP measures. Despite this there may be some residual confounding which we were not able to address.

The ALSPAC cohort data allowed us to incorporate wide range of measures of education and employment participation and level. However, these items are self-reported allowing the possibility of mis-reporting. We did not consider separately the role of cohabitation and parenthood in our analysis, as we wanted to focus primarily on socioeconomic trajectories. Previous research from older cohorts has suggested that early transitions to parenthood contribute to poor metabolic health, although this may not be completely independent of SEP [49,50]. Overall the proportion of the ALSPAC sample who had become parents by age 24y in the ALSPAC cohort was low, although highest in the Part-time Employment class (at 33% of females), which could have contributed in part to poor metabolic outcomes seen in this class.

## Conclusions

This analysis revealed a diversity of early adulthood socioeconomic trajectories, compared with earlier cohorts, in particular identifying a Part-time Employment trajectory, not seen in previous literature. Differences in adiposity and metabolic outcomes could be seen between classes, with those entering employment at age 18, and those in part-time employment identified as groups already showing evidence of higher risk for later cardiovascular disease by age 24y. More research is needed focussing on the latter population group and the factors contributing to the development of higher cardiovascular risk in this population during early adulthood.

## Acknowledgements

We are extremely grateful to all the families who took part in this study, the midwives for their help in recruiting them, and the whole ALSPAC team, which includes interviewers, computer and laboratory technicians, clerical workers, research scientists, volunteers, managers, receptionists and nurses.

## Funding

The UK Medical Research Council and Wellcome (Grant ref: 217065/Z/19/Z) and the University of Bristol provide core support for ALSPAC. This publication is the work of the authors and Eleanor Winpenny will serve as guarantor for the contents of this paper. A comprehensive list of grants funding is available on the ALSPAC website (http://www.bristol.ac.uk/alspac/external/documents/grant-acknowledgements.pdf); This research was specifically funded by the Wellcome Trust, UK Medical Research Council, British Heart Foundation including grant numbers 076467/Z/05/Z, CS/15/6/31468, MR/M006727/1, 076467/Z/05/Z, PG/06/145, 086676/7/08/Z, CS/15/6/31468, SP/F/21/150020, 102215/2/13/2, 102215/Z/13/Z, 092731, 092731, and G0401540 73080. EW is funded by the UK Medical Research Council: MR/T010576/1.

## Competing Interests

The authors have no relevant financial or non-financial interests to disclose.

## Author contributions

EMW designed the study, performed the analyses and drafted the manuscript. KT and LH advised on study design and statistical analysis, JS advised on latent class analysis in MPlus, AH advised on cardiometabolic outcomes. KT and LH commented on the analysis plans, and all authors contributed to the interpretation of results, made comments and edits to the manuscript, and read and approved the final manuscript.

## Ethics approval

Ethical approval for the study was obtained from the ALSPAC Law and Ethics Committee and the Local Research Ethics Committees, and conformed to the Declaration of Helsinki.

## Consent to participate

Consent for biological samples has been collected in accordance with the Human Tissue Act (2004). Informed consent for the use of data collected via questionnaires and clinics was obtained from participants following the recommendations of the ALSPAC Ethics and Law Committee at the time. If the child was younger than age 16 years, informed written consent was obtained from the parent/guardian alongside assent from the child. When participants were age 16 years or older, they provided their own informed written consent.

## Data and Code availability

The ALSPAC study website contains details of all the data that is available through a fully searchable data dictionary and variable search tool: http://www.bristol.ac.uk/alspac/researchers/our-data/.

Analysis code for the main analysis is found in the supplementary material, and further code requested from the corresponding author.

## Notes

### Competing Interest Statement

The authors have declared no competing interest.

### Author Declarations

The ALSPAC Law and Ethics Committee and the Local Research Ethics Committees gave ethical approval for this work.

