## Supplementary methods and tables for "How do socioeconomic trajectories experienced during early adulthood contribute to the development of cardiometabolic health in young adults?"

Mplus code for association of SEP class and outcomes: Step 3 of manual BCH method as described in Asparouhov and Muthen (2021).

```
TITLE: Latent ALSPAC SEP - using all outcomes data
DATA:
File = "V:\Behavioural_Epidemiology\Eleanor Winpenny\ALSPAC_SES\EA traj\Analysis\MPlus\
  Final BCH\Step3\By sex\Female\fullimplist.dat";
Type=Imputation;

VARIABLE:
Names =
id sex ethnic parity matage mated_16 IMD_16 famstr_8 alc_16 smok_16 cann_16
SRHLT_16 mfq_16 sysBP_15 bfat_15 parnt_24 zMetZ_24  waist_24 bfat_24 sysBP_24
diaBP_24 pulse_24 Trig_24 HDL_24 Gluc_24 HOMA_24 IMT_24 PWV_24 LVMI_24 RWT_24 CRP_24
BCHW1 BCHW2 BCHW3 BCHW4
MetZ_dat wai_dat bfat_dat sysBP_dat diaBP_dat puls_dat HOMA_dat Trig_dat
HDL_dat Gluc_dat IMT_dat PWV_dat LVMI_dat RWT_dat CRP_dat;
Missing = all (-9999) ;
IDVARIABLE = id;
USEVARIABLES =
 bfat_24 ETHNIC parity matage MATED_16 IMD_16 FAMSTR_8 ALC_16 SMOK_16 CANN_16
 SRHLT_16 MFQ_16 SYSBP_15 bfat_15 BCHW1 BCHW2 BCHW3 BCHW4;
USEOBSERVATIONS ARE (bfat_dat==1);
CLASSES=c(4);
TRAINING = bchw1-bchw4(bch);

define: center parity matage MATED_16 IMD_16 FAMSTR_8 ALC_16 SMOK_16 CANN_16
SRHLT_16 MFQ_16 SYSBP_15 bfat_15 (grandmean);
```

```
ANALYSIS:

TYPE=MIXTURE;

Starts=0;

Estimator=mlr;

MODEL:

%overall%

bfat_24 on ethnic parity matage mated_16 IMD_16 famstr_8 alc_16 smok_16 cann_16

SRhlt_16 mfq_16 sysBP_15 bfat_15;

C on ethnic parity matage mated_16 IMD_16 famstr_8 alc_16 smok_16 cann_16

SRhlt_16 mfq_16 sysBP_15 bfat_15;

%C#1%
[bfat_24] (MZ1);

bfat_24;

%C#2%

[bfat_24] (MZ2);

bfat_24;

%C#3%

[bfat_24] (MZ3);

bfat_24;

%C#4%

[bfat_24] (MZ4);

bfat_24;

!to test differences between individual pairs of classes

Model Constraint:

New (MZ1v2 MZ1v3 MZ1v4 MZ2v3 MZ2v4 MZ3v4);

MZ1v2 = MZ1-MZ2;

MZ1v3 = MZ1-MZ3;
```

```
MZ1v4 = MZ1-MZ4;
MZ2v3 = MZ2-MZ3;
MZ2v4 = MZ2-MZ4;
MZ3v4 = MZ3-MZ4;

!to test overall differences between classes
Model test:
  0=MZ1-MZ4;
  0=MZ2-MZ4;
  0=MZ3-MZ4;

 Output: Tech1 tech4 svalues sampstat;
```

Stata code for multiple imputation

A single multiple imputation was conducted, including all outcomes, BCH weights reflecting the class exposure, covariates, and auxiliary variables as shown in Supplementary Table S2. Imputation was conducted separately for males and females, in line with the sex-stratified analysis.

```
mi set mlong

mi register imputed ethnic parity matage  matedu_16 IMD_16 familystruc_8 alcohol_16 smoking_16 cannabis_16 partner_sec_18w partner_sec_32w partner_sec_4 matedu_8 matedu_32w IMD_14 IMD_8 famincome_4 famincome_8 SRhealth_16 mfq_16 systBP_15 bodyfat_15 parent_24 bodyfat_24 sysBP_24 diasBP_24 pulseBP_24 logTrig_24 HDL_24 Glucose_24 logHOMA IMT_24 PWV_24 LVMI_24 RWT_24 logCRP_24

 mi impute chained (logit) ethnic familystruc_8 parent_24 (ologit)  parity matedu_16 IMD_16 alcohol_16 smoking_16 cannabis_16 matedu_32w matedu_8 IMD_8 IMD_14 famincome_8 famincome_4 partner_sec_18w partner_sec_32w partner_sec_4 ///
(regress) matage SRhealth_16 mfq_16 systBP_15 bodyfat_15 bodyfat_24 sysBP_24 diasBP_24 pulseBP_24 logTrig_24 HDL_24 Glucose_24 logHOMA IMT_24 PWV_24 LVMI_24 RWT_24 logCRP_24 ///
 = BCHW1 BCHW2 BCHW3 , add(100) rseed(1234) dots augment

save imp_full_v4male.dta, replace
```

**Supplementary Table S1: Variables used in multiple imputation**

| Covariate | Type of variable | Model used to predict missing data in this variable | N (% of analysis dataset) with data on this variable |
|---|---|---|---|
| **Outcome variables** | | | |
| Body fat percentage, 24y | Continuous | Linear regression | 3846 (51%) |
| SBP, 24y | Continuous | Linear regression | 3951 (52%) |
| DBP, 24y | Continuous | Linear regression | 3951 (52%) |
| Heart rate, 24y | Continuous | Linear regression | 3951 (52%) |
| Log Triglycerides, 24y | Continuous | Linear regression | 3212 (43%) |
| HDL cholesterol, 24y | Continuous | Linear regression | 3214 (43%) |
| Log HOMA-IR, 24y | Continuous | Linear regression | 3214 (43%) |
| Glucose, 24y | Continuous | Linear regression | 3213 (43%) |
| Log CRP, 24y | Continuous | Linear regression | 2869 (38%) |
| cIMT, 24y | Continuous | Linear regression | 1955 (26%) |
| PWV, 24y | Continuous | Linear regression | 2284 (30%) |
| LVMI, 24y | Continuous | Linear regression | 1953 (26%) |
| RWT, 24y | Continuous | Linear regression | 1963 (26%) |
| **Exposure variables** | | | |
| BCH weights (BCHW1 BCHW2 BCHW3), reflecting the probability of classification of the individual in each class. | Continuous | n/a | 7526 (100%) |
| **Covariates** | | | |
| Sex | Binary | n/a | 7526 (100%) |
| Ethnicity | Binary | Logistic regression | 6742 (90%) |
| Maternal age during pregnancy | Continuous | Linear regression | 6815 (91%) |
| Maternal parity | Ordered Categorical | Ordered logistic regression | 6793 (90%) |
| Maternal education, age 16 | Ordered Categorical | Ordered logistic regression | 4374 (58%) |
| Neighbourhood deprivation, age 16 | Ordered Categorical | Ordered logistic regression | 4907 (65%) |
| Family structure, age 8 | Binary | Logistic regression | 5652 (75%) |
| Alcohol intake, 16y | Ordered Categorical | Ordered logistic regression | 5027 (67%) |
| Smoking, 16y | Ordered Categorical | Ordered logistic regression | 5034 (67%) |
| Cannabis use, 16y | Ordered Categorical | Ordered logistic regression | 5031 (67%) |
| Self-rated health 16y | Ordered Categorical | Linear regression | 5051 (67%) |
| Depressive symptoms 16y | Continuous | Linear regression | 5052 (67%) |
| Body fat %, 15y | Continuous | Linear regression | 4625 (61%) |
| Systolic blood pressure, 15y | Continuous | Linear regression | 4556 (61%) |
| **Auxiliary variables** | | | |
| Maternal education, 32 weeks | Ordered categorical | Ordered logistic regression | 6765 (90%) |
| Maternal education, 8y | Ordered categorical | Ordered logistic regression | 5617 (75%) |

| | | | |
|---|---|---|---|
| IMD, 8y | Ordered categorical | Ordered logistic regression | 5735 (76%) |
| IMD, 14y | Ordered categorical | Ordered logistic regression | 5047 (67%) |
| Family income, 4y | Ordered categorical | Ordered logistic regression | 5559 (74%) |
| Family income, 8y | Ordered categorical | Ordered logistic regression | 5161 (69%) |
| Partner occupational social class reported at age 18 weeks (partner report) | Ordered categorical | Ordered logistic regression | 4563 (61%) |
| Partner occupational social class, 32 weeks (mother report) | Ordered categorical | Ordered logistic regression | 5043 (67%) |
| Partner occupational social class, 4y (mother report) | Ordered categorical | Ordered logistic regression | 4090 (54%) |
| **Variable for supplementary analysis** | | | |
| Parenthood, 24y | Binary | Logistic regression | 6281 (83%) |

**Supplementary Table S2:** Fit indices for latent class models, testing different numbers of latent classes.

| n=7568 | | | | | | | | |
|---|---|---|---|---|---|---|---|---|
| Classes | Loglikeli-hood | AIC | BIC | aBIC | VLMR LRT (p-value) | BLRT (p-value) | entropy | class proportions (% of total population) |
| 2 | -61985.5 | 124297 | 125426.9 | 124908.9 | 0.00 | 0.00 | 0.573 | 39, 61 |
| 3 | -60155.6 | 120755 | 122293 | 121588 | 0.00 | 0.00 | 0.599 | 27, 36, 37 |
| 4 | -59291.1 | 119144.1 | 121092 | 120199 | 0.00 | 0.00 | 0.616 | 34, 23, 12, 30 |
| 5 | -58595.3 | 117870.5 | 120227.3 | 119146.9 | 0.2549 | 0.00 | 0.616 | 10, 23, 28, 25, 14 |
| 6 | -58158.5 | 117114.9 | 119880.7 | 118612.7 | 0.632 | 0.00 | 0.617 | 27,9,9,18,24,13 |

**Supplementary Results**

**Table S3: Estimated means (with 95% confidence intervals) of each outcome for each socioeconomic trajectory class, unadjusted for covariates**

| | **Male** | | | | | **Female** | | | | |
|---|---|---|---|---|---|---|---|---|---|---|
| | **Higher Education (class 1)** | **Late Education (class 3)** | **Part-time Employment (class 2)** | **Early Employment (class 4)** | **Wald test, p-value** | **Higher Education (class 1)** | **Late Education (class 3)** | **Part-time Employment (class 2)** | **Early Employment (class 4)** | **Wald test, p-value** |
| **Body fat percentage, % (n=3846)** | 23.8 (22.9,24.7) | 23.6 (22.5,24.7) | 26.1 (24.5,27.7) | 25.8 (24.9,26.8) | 0.002 | 33.8 (33.1,34.6) | 33.5 (32.6,34.4) | 36.7 (35.7,37.7) | 37.0 (36.2,37.8) | <0.001 |
| **Systolic blood pressure (n=3951)** | 123.0 (121.8,124.3) | 121.9 (120.3,123.6) | 122.3 (120.3,124.3) | 124.5 (123.1,125.9) | 0.105 | 112.0 (111.0,112.9) | 110.8 (109.7,111.9) | 111.4 (110.2,112.6) | 112.3 (111.3,113.3) | 0.206 |
| **Diastolic blood pressure (n=3951)** | 67.0 (66.1,68.0) | 67.1 (65.9,68.2) | 68.8 (67.1,70.4) | 68.5 (67.4,69.6) | 0.099 | 66.4 (65.7,67.2) | 65.9 (65.0,66.7) | 66.1 (65.2,67.1) | 67.2 (66.4,68.1) | 0.131 |
| **Heart rate, bpm (n=3951)** | 63.3 (62.0,64.5) | 63.8 (62.4,65.3) | 67.4 (65.4,69.3) | 64.5 (63.1,65.8) | 0.014 | 67.1 (66.2,68.1) | 69.0 (67.7,70.2) | 69.9 (68.7,71.1) | 69.3 (68.3,70.3) | 0.006 |
| **Triglycerides, mmol/L (n=3212)** | 0.92 (0.87,0.98) | 0.90 (0.84,0.96) | 1.04 (0.95,1.14) | 0.97 (0.91,1.04) | 0.061 | 0.81 (0.78,0.85) | 0.80 (0.76,0.84) | 0.87 (0.82,0.92) | 0.87 (0.83,0.91) | 0.021 |
| **HDL cholesterol, mmol/L (n=3214)** | 1.46 (1.42,1.51) | 1.46 (1.40,1.53) | 1.30 (1.24,1.37) | 1.32 (1.28,1.37) | <0.001 | 1.73 (1.68,1.78) | 1.70 (1.65,1.75) | 1.54 (1.47,1.60) | 1.58 (1.53,1.63) | <0.001 |
| **HOMA-IR (n=3214)** | 0.773 (0.713,0.837) | 0.766 (0.698,0.842) | 0.967 (0.844,1.107) | 0.871 (0.791,0.959) | 0.016 | 0.861 (0.807,0.918) | 0.857 (0.793,0.927) | 0.961 (0.873,1.058) | 0.967 (0.899,1.039) | 0.042 |
| **Glucose, mmol/L (n=3213)** | 5.44 (5.36,5.51) | 5.48 (5.34,5.62) | 5.40 (5.29,5.50) | 5.45 (5.37,5.53) | 0.857 | 5.19 (5.12,5.26) | 5.17 (5.11,5.23) | 5.17 (5.10,5.24) | 5.20 (5.12,5.27) | 0.941 |
| **C-Reactive Protein, mg/L (n=2869)** | 0.712 (0.621,0.817) | 0.605 (0.512,0.715) | 0.682 (0.548,0.850) | 0.895 (0.768,1.043) | 0.008 | 0.965 (0.851,1.094) | 0.841 (0.729,0.971) | 0.966 (0.808,1.154) | 1.126 (0.980,1.295) | 0.041 |
| **Carotid Intima media thickness, mm (n= 1955)** | 0.470 (0.460,0.480) | 0.467 (0.455,0.479) | 0.465 (0.453,0.477) | 0.464 (0.454,0.474) | 0.891 | 0.453 (0.447,0.459) | 0.454 (0.446,0.462) | 0.459 (0.451,0.467) | 0.449 (0.441,0.457) | 0.376 |

| | | | | | | | | | | |
|---|---|---|---|---|---|---|---|---|---|---|
| **Carotid-femoral pulse wave velocity, m/s (n=2284)** | 6.77 (6.58,6.96) | 6.72 (6.47,6.97) | 6.46 (6.20,6.72) | 6.63 (6.45,6.80) | 0.336 | 6.09 (5.96,6.21) | 6.06 (5.88,6.23) | 6.07 (5.90,6.24) | 6.10 (5.96,6.24) | 0.987 |
| **Left ventricular mass index, g/m$^{1.7}$ (n=1953)** | 59.8 (57.5,62.1) | 56.7 (54.3,59.1) | 55.4 (52.6,58.1) | 60.2 (57.7,62.7) | 0.027 | 47.9 (46.6,49.2) | 47.5 (45.9,49.1) | 48.9 (46.8,51.0) | 49.6 (47.8,51.4) | 0.307 |
| **Relative wall thickness (n=1963)** | 0.372 (0.362,0.382) | 0.363 (0.351,0.375) | 0.368 (0.352,0.384) | 0.371 (0.359,0.383) | 0.721 | 0.369 (0.361,0.377) | 0.368 (0.356,0.380) | 0.351 (0.339,0.363) | 0.364 (0.354,0.374) | 0.113 |

Geometric means and confidence intervals are reported for Triglycerides, CRP and HOMA-IR, and Wald tests for differences between classes are reported for the log-transformed outcomes for Triglycerides, CRP and HOMA-IR.

**Supplementary Table S4: Estimated means (with 95% confidence intervals) of Relative Wall Thickness for each socioeconomic trajectory class, adjusted for covariates, and restricted to females without children by age 24y.**

| | **Higher Education** | **Late Education** | **Part-time Employment** | **Early Employment** | **Wald test, p-value** |
|---|---|---|---|---|---|
| **Relative wall thickness (n=1,077)** | 0.370 (0.360,0.380) | 0.374 (0.328,0.360) | 0.344 (0.328,0.360) | 0.359 (0.347,0.371) | 0.0337 |
